# Pre-Diagnostic Circulating RNAs Networks Identify Testicular Germ Cell Tumour Susceptibility Genes

**DOI:** 10.1101/2022.12.16.22283563

**Authors:** Joshua Burton, Trine B. Rounge, Trine B. Haugen, Marcin W. Wojewodzic

**Affiliations:** Department of Life Sciences and Health, OsloMet – Oslo Metropolitan University, Oslo, Norway; Medical Research Council Integrative Epidemiology Unit, University of Bristol, Bristol, UK; Centre for Bioinformatics, Department of Pharmacy, University of Oslo, Norway; Department of Research, Cancer Registry of Norway, Norway; Department of Environment and Health, Norwegian Institute of Public Health, Oslo, Norway

**Keywords:** Network analysis, pre-diagnostic, TGCT, testicular cancer, RNA

## Abstract

Testicular germ cell tumour (TGCT) is a malignancy with known inherited risk factors, affecting young men. We have previously identified several hundred circulating RNAs that were differentially expressed in pre-diagnostic serum samples from TGCT cases when compared to healthy controls. In this study we performed network preservation analyses of pre-diagnostic serum mRNA and miRNA. Hub genes, enriched functional pathways, and regulatory feature prediction were identified for all TGCT, seminoma, and non-seminoma cases separately, compared to controls. We identified *UBCA1, RCC1, FMR1, OSA3,* and *UBE2W* as hub genes associated with TGCT. The genes *OSA3* and *UBE2W* have previously been associated with testicular dysgenesis syndrome (TDS) disorders. Previously described TGCT susceptibility genes *TEX14*, *NARS2,* and *G3BP2* were identified as hub genes in both seminoma and non-seminoma networks. Furthermore, network module analysis showed prediction of transcription factors for oestrogen-related receptors. The overlap between network hub genes and TGCT susceptibility genes indicates a role in the progression from germ cell neoplasia in situ (GCNIS) to TGCT that should be further investigated.

## Introduction

Testicular germ cell tumour (TGCT) is the most common form of testicular cancer affecting primarily younger males. Throughout the 20^th^ and 21^st^ century there have been rising rates of TGCT in developed countries. However, in recent years the incidence rates have begun to plateau in Northern and Western European countries with high incidence, in Norway at around 11 cases per 100,000 person-years (1–3). It is generally agreed that TGCT develop from a pre-malignancy, from germ cell neoplasia in situ (GCNIS), obtained through a loss of differentiation of gonocytes during foetal life which progress to a malignancy during postnatal puberty (4). There are around 80 risk loci for TGCT as well as environmental risk factors, and ethnicity also affects the likelihood of developing TGCT (5–8). Further disorders commonly associated with the testicular dysgenesis syndrome (TDS) include cryptorchidism, poor semen quality, and hypospadias (9). These disorders are known to be associated with higher risk of TGCT to varying degrees (10). However, the aetiology behind TGCT is still mainly unknown.

Advances in TGCT treatment have led to an increased 5-year survival rate, up to 95% (11), however, survivors of TGCT are still at risk of secondary cancers, cardiovascular issues, epigenetic alterations, and reduced longevity associated with cisplatin treatment (12–15). It is therefore important to diagnose TGCT early to reduce the rounds and dosage of cisplatin-based treatment needed and therefore potentially reduce the long-term side effects of cisplatin. Novel methods of analysing existing data are therefore needed.

Analyses of pre-diagnostic samples may increase understanding of the progression of TGCT. The pre-diagnostic serum samples from the Janus Serum Bank are well suited for such studies (16, 17). Our previous studies of pre-diagnostic samples from this biobank using traditional linear models with negative binomial distribution employed on count data, identified genes associated with the development of lung and TGCT (18, 19). A total of 818 circulating RNAs were found to be differentially expressed in pre-diagnostic TGCT cases when compared to controls. For instance, differences in expression were observed for the male fertility-related gene, *TEX101*, and the X-chromosome inactivation gene, *TSIX* (18).

Weighted gene correlation network analysis (WGCNA) is a systems biology approach that allows for increased comprehension of large and multidimensional transcriptomic datasets (20). WGCNA identifies gene co-expression patterns between multiple samples from various states. Gene co-expression can be used to identify a variety of elements such as future therapeutic targets or biomarkers as well as screening datasets for networks of genes relating to certain traits (21). WGCNA also allows for the comparison of such networks to show nodes and clusters that are preserved between differing states, such as histological subtypes. This is one of the major differences between standard RNA-Seq approaches, where linear models are deployed (limma, edgeR, or DESeq2) (22–24). WGCNA does not only consider the differentially expressed genes, instead opting to utilise the comprehensive gene list to search for inter-connectivity between clusters of genes (20). Using these methods, previous research has found related genes to specific cancers through mRNA and miRNA analyses, including lung adenocarcinoma (25), bladder cancer (26), and breast cancer (27).

The WGCNA can be used to obtain new information about the mechanisms underlying TGCT development. Alterations in the networks may potentially indicate the presence of a disease or precede the development of one (28, 29). Observed changes within gene networks also have the potential to be used for biomarker discovery (30). Extensive networks relying on multiple different biological components, such as transcription factors, mRNAs, miRNAs, miRNA targets, and have been demonstrated in multiple diseases, including TGCT development (31). The data in Mallik’s work originated from post-diagnostic samples of seminoma and non-seminomas and compared only TGCT cases without controls. Subtype-specific networks were constructed, and mRNAs, miRNA and TFs were identified. In both the seminoma and non-seminoma modules, the top miRNAs were onco-related miRNAs for several types of cancers. Common hub genes in Mallik’s work, *SP1* and *MYC,* were also identified between the networks constructed for seminomas and non-seminomas.

Post-diagnostic TGCT networks contains Phosphatase and Tensin Homolog (*PTEN*), targeted by three miRNAs, hsa-miR-141, hsa-miR-222, and hsa-miR-21, with hsa-miR-222 also regulating KIT and Tumor Protein 53 (*TP53*) (32). The complexity of the networks can often be overlooked when only differential expression is studied, therefore a more in-depth method of comparing disease states should be pursued in order to obtain a more complete understanding of what drives TGCT development.

We aimed to identify mRNA and miRNA networks associated with TGCT development in pre-diagnostic serum samples by using WGCNA. We compared networks from different TGCT histological types. We hypothesised that by using network analysis, we could propose genes and mechanisms related to TGCT development.

## Materials and Methods

### Data and sample description

We proceeded with the data in our previous study of pre-diagnostic RNA profile in serum from TGCT patients (18). In brief, we obtained samples from the Janus Serum Bank that were linked to registry data from the Cancer Registry of Norway to identify matched cases and controls. We included patients who developed TGCT up to 10 years after sample donation and control samples who were cancer-free within 10 years after sample donation. The serum samples were collected between 1972 – 2004 from participants in health surveys and red cross blood donors.

In total, 79 TGCT cases were identified that had blood donated up to 10 years prior to TGCT analyses. Of these, 52 were identified as seminoma, and 27 as non-seminoma. In addition, 111 matched controls that were cancer-free 10 years after blood donation were also retrieved. This sample set is part of a larger data collection named JanusRNA, described in detail by Langseth., et al (33). RNA profiles were generated by extracting RNAs from 400 µl serum, and small RNA sequencing libraries were produced with NEBNext kit (Cat. No E7300, New England Biolabs Inc.) and sequenced on the HiSeq 2500 (Illumina) as previously described (34).

### Bioinformatics analysis

Pre-processing of raw transcriptomics data was run using a high-performance computing cluster at the Cancer Registry of Norway. Detailed description can be found in a previously published article (34) (https://github.com/sinanugur/sncRNA-workflow). In brief, the reads generated an average depth of 18.4 million raw reads per sample. Adapters were removed with AdapterRemoval (v.2.1.7) with collapsed reads then being mapped to the human genome (hg38) with Bowtie2 (v2.2.9). Annotation set for RNAs was GENCODE (v.26) and for miRNAs was miRbase (v.22). We included mRNAs with at least five reads in more than 20% of the samples, and in the miRNA analysis we removed miRNAs that had no reads in more than 10% of the samples.

Controls and cases for the network construction were matched using age at sampling, time of sampling, and blood donation group, including separate matching for samples in the ‘0-7 years before diagnosis’ and ‘0-4 years before diagnosis’ groups as part of a sensitivity analysis (Figure 1 I). The same matching methods were performed between the subtypes, seminomas and non-seminomas during the network analysis based on TGCT histology. The *optmatch* R package (v0.9–11) (github.com/markmfredrickson/optmatch) was used to find these matching sets. Full overview of these procedures was given in a previous article (Burton et al., 2020).

**Figure 1.**
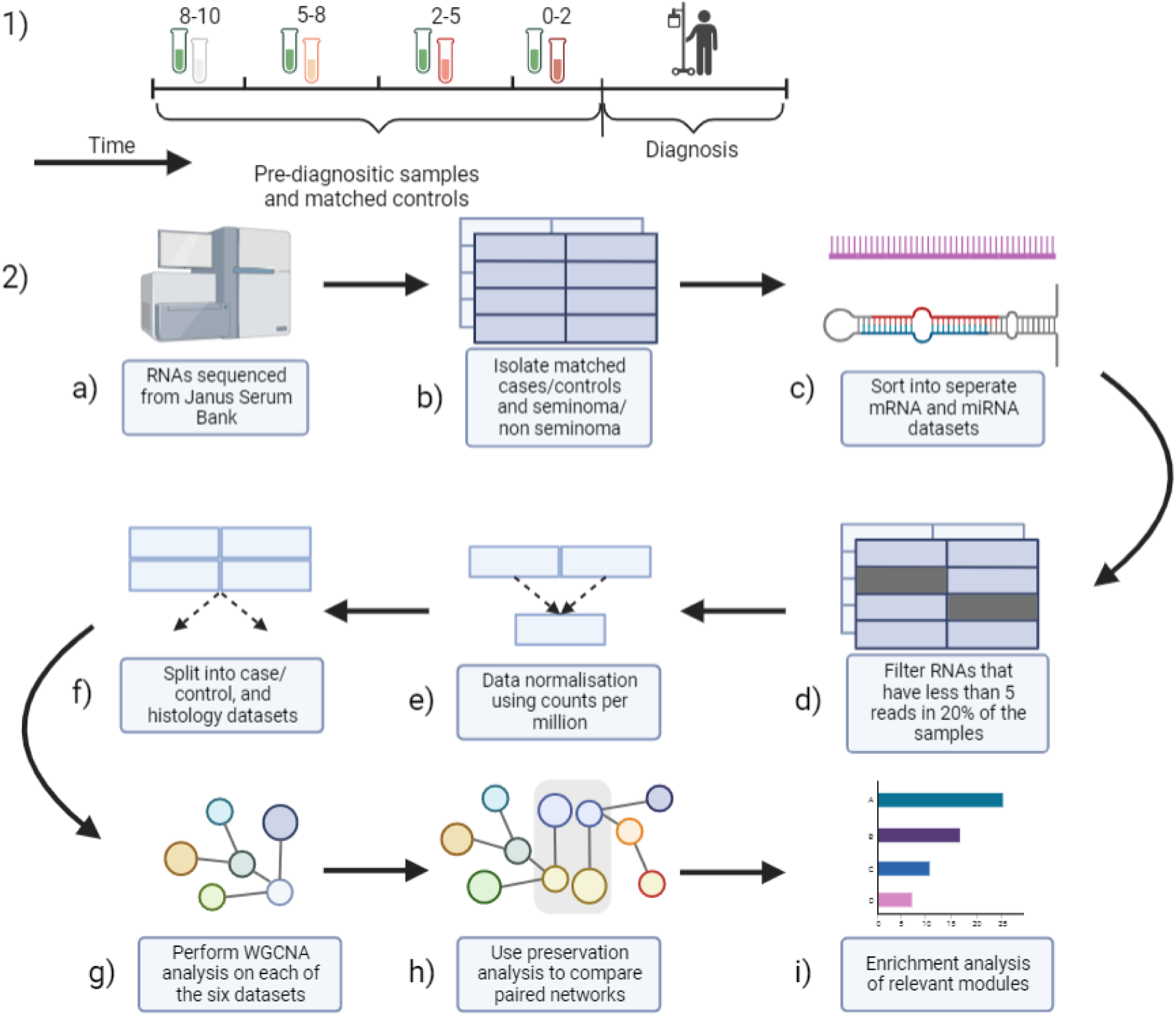
I) Design of the study. Samples were identified from the Janus Serum Bank (JSB) in periods prior to diagnosis. II) Workflow for analysis. a) RNAs were initially sequenced from JSB serum sample, and sncRNAs were quantified. b) Samples were split based on cancer diagnosis and histology and matched with controls. c) mRNA and miRNA datasets were separated for different analyses. d, e & f) Quality control was performed on the datasets, and data was prepared for network analyses. g) WGCNA pipelines were used to perform network analyses. h) The preservation analyses were then performed on comparable networks. i) Enrichment analysis was then performed on relevant modules.

Statistical analyses were performed on the processed datasets. Firstly, counts were normalised using counts per million from the EdgeR package (22). The R package ComBat (35) was then used to determine if there was any batch effect in our data. A simplified WGCNA analysis on mRNA cases data which had undergone batch correction was run. Power graph analyses were compared with the full mRNA cases analysis which had not undergone batch correction.

### WGCNA pipeline steps to module preservation analysis

Weighted Gene Correlation Network Analysis (WGCNA) package (1.70-3) was used on the pre-processed data. First, we ensured scale free topology that is necessary assumption for network analysis by choosing an appropriate power value for all analyses. WGCNA was then run through the following steps described in the flowchart (Figure 1 II). Firstly, feature (mRNA or miRNA expression) correlations were used to construct the individual networks, followed by clusters identification of interconnected nodes hereafter referred to as modules. Then the nodes within the modules are summarised through a highly connected hub node (‘eigengenes’ or ‘eigenmirs’ for mRNA and miRNA network, respectively), alleviating multiple testing issues by reducing the dimensions of the datasets. The distinct individual modules, named by colours, can then be identified, and all nodes within the network can be annotated depending on how close they are to the modules. In addition, TGCT samples were divided by seminoma and non-seminoma samples, and network construction was performed as above, with only mRNA counts. This was due to the lack of sufficient miRNAs needed for network analysis in each histology.

### Preservation network analysis

Network preservation analysis was used for the comparison of two networks and the modules preserved across networks. We identified modules that are preserved across the two constructed networks and the degree to which they are preserved. For the network preservation analysis, we made several comparisons between groups, initially, contrasts between mRNA serum levels in cases and controls, followed by miRNA serum levels between cases and controls and mRNA serum levels between seminomas and non-seminomas. For each of these subsets we constructed separate networks, whilst also assuring the same scale-free topology. Module preservation statistics were computed using the modulePreservation function (200 permutations) implemented in WGCNA (36) and further implemented from Frisch’s work, (37). Network module preservation statistics quantify how density and connectivity patterns of modules defined in a reference dataset (control samples) are preserved in a test dataset (TGCT samples). Network adjacency comparisons are superior estimates of module preservation to standard cross-tabulation techniques (36). We used network adjacency comparison to assess preservation of gene co-expression patterns in control and TGCT modules, and between histology types. The overall significance of the preservation was assessed using Zsummary and median rank statistics (36). Based on the thresholds proposed in the documentation, Zsummary <2 indicates no preservation, 2 < Zsummary < 10 weak to moderate evidence of preservation, and Zsummary >10 strong evidence of module preservation across networks. Median rank statistics further consider module size for preservation assessment (36).

### Pathway analysis

Functional annotation was performed using the enrichment analysis tool Enrichr (38–40), by inputting gene lists from modules generated by WGCNA. Of particular interest were modules that were not preserved between cases and controls and those which were preserved between seminomas and non-seminomas. Enrichr databases were then reviewed for significantly enriched pathways and pathways associated with TGCT or TDS conditions.

### Motive finder, transcription factor mRNA

The regulatory patterns of the mRNAs included in the modules were investigated using a promoter motif analysis tool i-cisTarget (https://gbiomed.kuleuven.be/apps/lcb/i-cisTarget/ Last accessed 6th June 2022). A full analysis with standard parameters was run with gene symbols as the input and RefSeq r45 and v.6 of the i-cisTarget database, with only TF binding site databases included in the analysis. This analysis could reveal if the module is regulated by specific transcription factors for genes related to male fertility or cancer development.

### Determining miRNA targets on mRNA

New modules predicting miRNA as preserved between cases and controls underwent a miRNA prediction analysis to determine which mRNAs the preserved miRNAs targeted. By translating miRNA to mRNA targets it also allows for easier comparison between the miRNA and mRNA networks. MiRNA targets are determined through sequence matching. Targets were extracted from miRDB (v5.0) with a cut off score of > 70 allowing only targets of high confidence to be included in the analysis.

### Enrichment analyses

Enrichment analyses were performed on the network and module data, firstly, KEGG analysis was performed, using Enrichr once more, on the top ten hub genes in each of the mRNA cases, seminoma and non-seminoma modules. A parallel enrichment analysis was also performed on all the genes in the module to give an overall prediction for enriched pathways. For miRNA cases, mRNA targets were used for enrichment analysis, with the cut off score used previously (>70).

### Cytoscape visualisation

Cytoscape (v. 3.9.0) was used to visualise the network modules. Cytoscape files were generated using the R package WGCNA’s export to Cytoscape function, generating both the node and the edge files necessary to visualise. Additional layout packages from yFiles Layout Algorithms (v. 1.1.2) were used for visualisation of the networks.

### Sensitivity analysis

In order to determine if the time to diagnosis of each sample affected network construction, a sensitivity analysis was performed by excluding the samples with a longer time period between sampling and diagnosis. As the data was split into four separate time frames from a previous study ([0-2], [2-5], [5-8], [8-10]), we first excluded the 8-10 years between sampling and diagnosis samples, which also gives us smaller datasets to work with. As a further sensitivity step, the 5-8 years between sampling and diagnosis samples were removed for a second round.

## Results

### Power calculations

To ensure scale-free topology, we performed power analysis and selected a power value equal to 10 for mRNA networks and miRNA networks (Sup F1).

#### Main characteristics of the networks

Network statistics showed that the mRNA cases network consisted of 19 modules, containing a total of 10596 mRNAs, with an average module density of 0.026 and an average heterogeneity of 0.99. The control network for mRNA consisted of 11 modules containing 8462 mRNAs, with an average module density of 0.028 and an average heterogeneity of 1.06.

The miRNA cases network consisted of five modules containing 403 miRNAs, with an average module density of 0.09 and average heterogeneity of 0.52, whilst the miRNA control network consisted of four modules containing a total of 403 miRNAs, with an average module density of 0.1 and an average heterogeneity of 0.49.

The seminoma network consisted of 22 modules containing 11032 mRNAs, with an average module density of 0.05 and average heterogeneity of 0.72, whilst the non-seminoma network consisted of 26 modules containing 11026 mRNAs, with an average module density of 0.06 and an average heterogeneity of 0.72.

### Modules of interest in mRNA cases vs. control

Modules with TGCT-related genes in mRNA cases include the black(cases), magenta(cases), purple(cases), salmon(cases), and the midnightblue(cases) modules (Table 5). The midnightblue(cases) module consisted of 169 genes with a density of 0.03, and the top three eigengenes for this module were *RTCB*, *JAKMIP1,* and *MAP3K12* (Table 1). The black(cases) module consisted of 580 genes with a density of 0.019, and the top three eigengenes in this module were *UBAC1*, *UBE2W* and *OAS3*. The magenta(cases) module consisted of 343 genes with a density of 0.025, and the top three eigengenes in this module were *BEST3, SLC16A2,* and *RCC1*. Purple(cases) module consisted of 303 genes with a density of 0.028, and the top three eigengenes in this module were *CPNE5*, *FMR1,* and *RBFOX2*. Salmon(cases) module consisted of only 194 genes with a density of 0.038, and the top three eigengenes in this module are *KIAA2026*, *IGF2BP3,* and *SBF2*.

**Table 1.** The top three eigengenes shared between the new midnightblue_(cases)_ module and the preserved red_(NS)_ module, with their gene name and gene ontology annotation. These modules were chosen as they shared the same eigengenes and were specific to TGCT cases.

| Gene ID | Gene Name | Gene Ontology Annotations |
| --- | --- | --- |
| *JAKMIP1* | Janus Kinase and Microtubule Interacting Protein 1 | RNA binding, kinase binding |
| *MAP3K12* | Mitogen-Activated Protein Kinase Kinase Kinase 12 | Protein homodimerization activity, protein kinase activity |
| *RTCB* | RNA 2',3'-Cyclic Phosphate and 5'-OH Ligase | RNA binding, RNA ligase (ATP) activity |

### mRNA cases vs. controls preservation analysis

Preservation analysis was performed on the mRNA case/control networks to see which modules were preserved and which were not. Modules that were present in cases but absent from controls, are called new modules, and in total there were 11 new modules in this analysis. New modules are of particular interest as they could show important genes involved in TGCT development. Modules where a significant number of mRNAs are shared between paired modules, are called preserved modules, seven in total. Finally, modules where several mRNAs are shared between paired modules but not to a significant degree, are known here as non-preserved modules, and there is only one.

Blue(cases)/blue(controls) showed significant preservation, with 603 preserved genes, and a p-value < 0.001. The case module midnightblue(cases) was identified as being a new module, as well as black(cases), magenta(cases), purple(cases_)_ and salmon(cases) modules.

### Modules of interest in Seminoma (SE) and Non-seminoma (NS)

The modules of interest for TGCT development in the SE network were the blue_(SE)_, lightyellow_(SE)_, and midnightblue_(SE)_. Blue_(SE)_ consists of 1251 genes, with a density of 0.044. The top three eigengenes for this module are *UBE2W*, *OAS3,* and *TNFAIP2*. Lightyellow_(SE)_ consists of 168 genes, with a density of 0.06. The top three eigengenes for this module are *UVSSA*, *KLHL21,* and *FIGNL1*. The module mightnightblue_(SE)_ consists of 231 genes, with a density of 0.097. The top three eigengenes for this module include *ZC4H2*, *ATP1B1,* and *GRID2*.

The modules of interest in the NS network were yellow_(NS)_, which consists of 947 genes, with a density of 0.032. The top three eigengenes for this module are *CROT*, *HELB,* and *PRELID3A*. The second module of interest was red_(NS)_, which consisted of 636 genes, with a density of 0.044. The top three hub genes for this module were *JAKMIP1*, *MAP3K12,* and *RTCB* (Table 1).

### SE vs. NS network’s preservation analysis

Preservation analysis of histology-related networks identified several modules of interest that were preserved between seminomas and non-seminomas networks. Preserved modules show commonalities between seminomas and non-seminomas. Of the 22 modules in the seminoma network and the 26 in the non-seminoma network, 20 were preserved between the two. One such module that was significantly preserved was the blue_(SE)/_red_(NS)_ module, which shared 114 genes between both histology states. There were also two new modules present for seminomas and six new modules present for non-seminomas, showing possible histology-specific genes of interest. Of note is the recurring presence of this module between all case networks.

### Modules of interest in miRNA cases

The module of interest within miRNA cases was the brown_(miCases)_ module, which consisted of 89 genes and had a density of 0.065. The top three eigenmiRs for this module were hsa-miR-30e-5p, hsa-miR-191-5p, and hsa-miR-199a-3p (Table 2).

**Table 2.** The top three eigenmiRs for the miRNA module brown_(miCases)_ with top ten highest confidence mRNA targets IDs.

| miRNA ID | Top 10 mRNA target IDs |
| --- | --- |
| hsa-miR-30e-5p | TWF1, B3GNT5, WDRZ, SCN2A, BRWD3, PTGFRN, DCUN1D3, NFAT5, KLHL20, PPARGC1B |
| hsa-miR-191-5p | NEURL4, TAF5, CREBBP, TMOD2, TJP1, MGST3, CBLN4, SPO11, INO80D, DDHD1 |
| hsa-miR-199a-3p | ADAMTSL3, KATNBL1, CELSR2, KLHL3, MAP3K4, ITGA3, LRP2, ETNK1, NID2, NAA25 |

### miRNA cases vs. controls preservation analysis

Preservation analysis of miRNA networks identified four modules that were preserved between cases and controls and one new module, specific to cases. mRNA target prediction for the top three eigenmiRs within this module was performed using > 70 as the cut off value for prediction score (Table 2). A total of 1310 unique mRNA targets were identified. EigenmiRs targeting prediction showed that all three eigengenes were predicted to target *MAP3K* genes as well as related pathway genes. Hsa-miR-30e-5p was predicted to target *KRAS* and other members of the RAS oncogene family, as well the gene *MAP3K12*, with a confidence score of 78.

### Enrichment Analyses

KEGG analysis of the mRNA TGCT cases network showed enrichment of the Ubiquitin mediated proteolysis pathway in lightcyan_(cases)_. Purple_(cases)_ showed significant enrichment of the Notch signalling pathway. Magenta_(cases)_, a new module, showed significant enrichment of the AMPK signalling pathway. Finally, the preserved green_(cases)_ module also showed significant enrichment of pathways in cancer (Table 3). Furthermore, the red_(SE)_ module showed significant enrichment of the T cell receptor signalling pathway (Table 4). Salmon_(NS)_, a preserved module between seminomas and non-seminomas, shows significant enrichment of the non-small cell lung cancer associated pathway (Table 5).

**Table 3.**
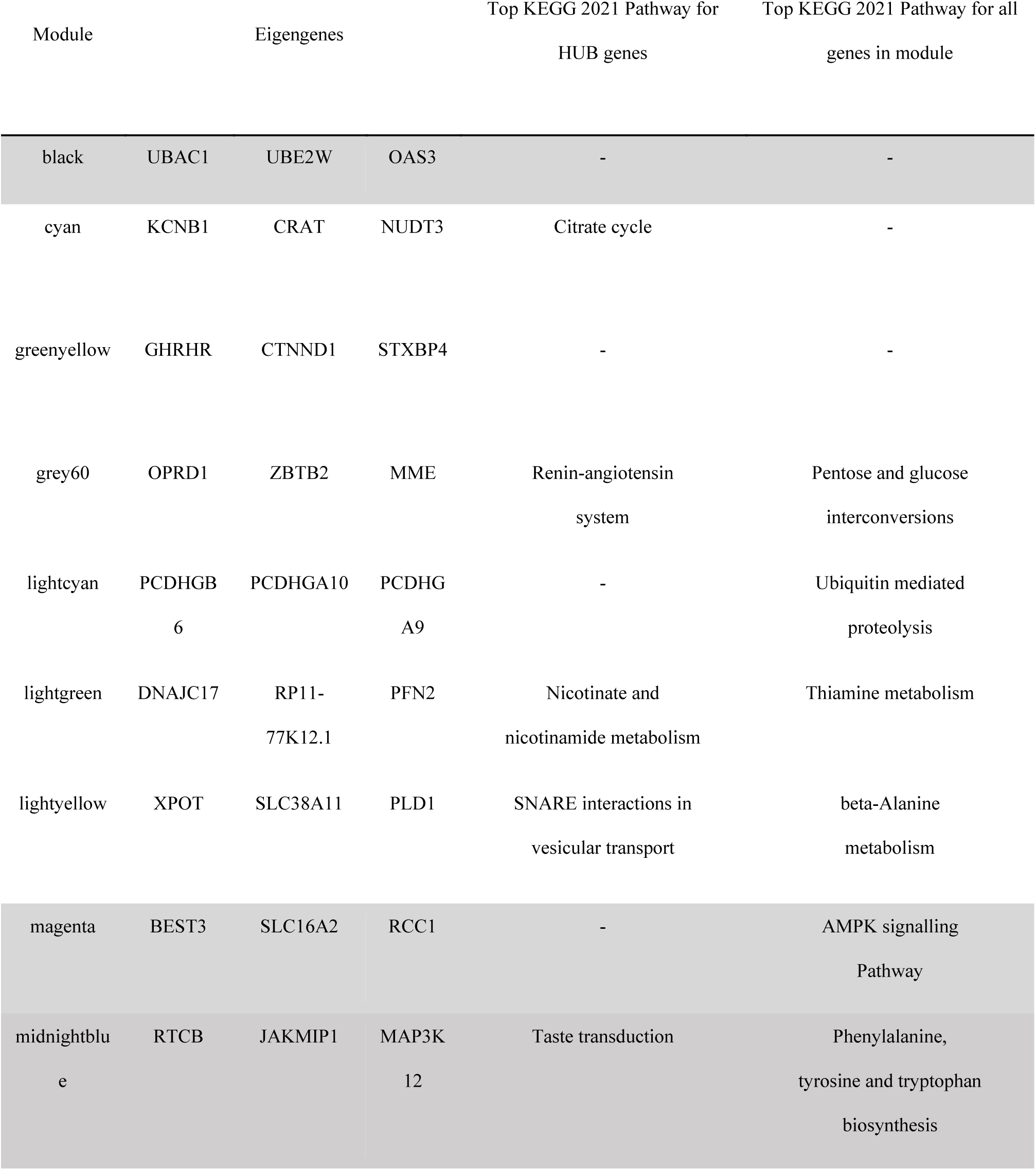
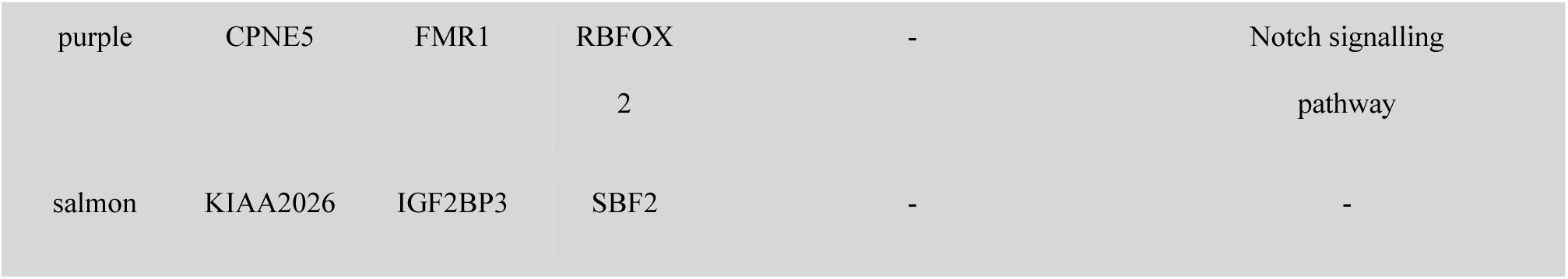
mRNA case network modules specific to mRNA cases when compared to mRNA controls. Top three eigengenes in each module listed, as well as significant enrichment analysis results for top 10 eigengenes and for all genes included in the module. Highlighted rows contain genes of interest within the top three eigengenes.

| Module | Eigengenes |  |  | Top KEGG 2021 Pathway for<br>HUB genes | Top KEGG 2021 Pathway for all<br>genes in module |
| --- | --- | --- | --- | --- | --- |
| black | UBAC1 | UBE2W | OAS3 | - | - |
| cyan | KCNB1 | CRAT | NUDT3 | Citrate cycle | - |
| greenyellow | GHRHR | CTNND1 | STXBP4 | - | - |
| grey60 | OPRD1 | ZBTB2 | MME | Renin-angiotensin<br>system | Pentose and glucose<br>interconversions |
| lightcyan | PCDHGB<br>6 | PCDHGA10 | PCDHG<br>A9 | - | Ubiquitin mediated<br>proteolysis |
| lightgreen | DNAJC17 | RP11-77K12.1 | PFN2 | Nicotinate and<br>nicotinamide metabolism | Thiamine metabolism |
| lightyellow | XPOT | SLC38A11 | PLD1 | SNARE interactions in<br>vesicular transport | beta-Alanine<br>metabolism |
| magenta | BEST3 | SLC16A2 | RCC1 | - | AMPK signalling<br>Pathway |
| midnightblue | RTCB | JAKMIP1 | MAP3K<br>12 | Taste transduction | Phenylalanine,<br>tyrosine and tryptophan<br>biosynthesis |
| purple | CPNE5 | FMR1 | RBFOX<br>2 | - | Notch signalling<br>pathway |
| salmon | KIAA2026 | IGF2BP3 | SBF2 | - | - |

**Table 4.** mRNA network modules for seminomas, with preservation state shows mRNA seminoma vs non-seminoma analysis results. Included are the top three eigengenes in each module as well as enrichment analysis results for top 10 eigengenes and for all genes included in the module. Modules that were preserved show number of genes preserved between seminomas and non-seminomas. Highlighted rows contain genes of interest within the eigengenes.

| Module | Eigengenes |  |  | Top KEGG 2021<br>Pathway for Top 10<br>HUB genes | Top KEGG 2021<br>Pathway for All<br>Genes in Module | Preservation state |
| --- | --- | --- | --- | --- | --- | --- |
| black | *USP14* | *GHRHR* | *CTNND1* | - | - | Preserved (21) |
| blue | *UBE2W* | *OAS3* | *TNFAIP2* | - | - | Preserved (114) |
| brown | *CFAP100* | *TTC7B* | *NRCAM* | - | - | Preserved (181) |
| cyan | *RP1-27O5.3* | *PSEN1* | *CERKL* | - | - | Preserved (26) |
| darkgreen | *NLRP11* | *DNAJB14* | *ADCY7* | - | - | Preserved (12) |
| darkred | *HEPH* | *HMG20B* | *ZNF565* | - | - | Preserved (6) |
| green | *DPPA2* | *PUDP* | *TMPRSS6* | - | Ascorbate and<br>aldarate<br>metabolism | Preserved (44) |
| greenyellow | *GLIPR1* | *TBX18* | *ITPKB* | - | - | Preserved (40) |
| grey60 | *FAM151B* | *EEA1* | *RAD23B* | - | - | Preserved (32) |
| lightcyan | *FASN* | *SCN2B* | *C1orf198* | - | - | Preserved (8) |
| lightgreen | *PPP1R13B* | *UBXN8* | *ZNF280C* | - | - | New |
| lightyellow | *UVSSA* | *KLHL21* | *FIGNL1* | - | - | New |
| magenta | *SCN5A* | *CLUH* | *ST8SIA1* | - | - | Preserved (115) |
| midnightblue | *ZC4H2* | *ATP1B1* | *GRID2* | - | - | Preserved (17) |
| pink | *TBX5* | *HSPA9* | *EIF4G1* | - | Shigellosis | Preserved (68) |
| purple | *RP11-729L2.2* | *SMAD4* | *PNPLA7* | Viral myocarditis | - | Preserved (11) |
| red | *BEST3* | *SLC16A2* | *CAMK1D* | - | T cell receptor signalling pathway | Preserved (151) |
| royalblue | *RAB8B* | *ZNF143* | *POM121C* | - | - | Preserved (10) |
| salmon | *SHC3* | *UGGT1* | *IARS2* | - | - | Preserved (9) |
| tan | *PDE6D* | *CENPN* | *LARP4* | - | - | Preserved (20) |
| turquoise | *CHRNA7* | *NECTIN3* | *PPP1R9A* | - | - | Preserved (549) |
| yellow | *PANK3* | *PANK2* | *COPZ1* | Pantothenate and CoA biosynthesis | Hippo signalling pathway | Preserved (173) |

**Table 5.** mRNA network modules for non-seminomas, with preservation state shows mRNA non-seminoma vs seminoma analysis results. Included are the top three hub genes in each module as well as enrichment analysis results for all 10 hub genes and for all genes included in the module. Modules that were preserved show number of genes preserved between seminomas and non-seminomas. Highlighted rows contain genes of interest within the eigengenes.

| Module | Eigengenes |  |  | Top KEGG<br>2021 Pathway<br>for Top 10 HUB<br>genes | Top KEGG 2021<br>Pathway for All Genes<br>in Module | Preservation state |
| --- | --- | --- | --- | --- | --- | --- |
| black | *CRAT* | *SLC6A9* | *KCNB1* | Glycosaminogly<br>can biosynthesis | - | Preserved (68) |
| blue | *LMBR1* | *NPFFR2* | *STPG2* | - | - | Preserved (549) |
| brown | *IL12RB1* | *HMGXB4* | *CHRNA1* | - | - | Preserved (40) |
| cyan | *NREP* | *GFRAL* | *ELMOD3* | - | - | New |
| darkgreen | *ALMS1* | *CWF19L2* | *KIF18A* | - | Ascorbate and aldarate<br>metabolism | Preserved (6) |
| darkgrey | *PRKACB* | *ANKRD13A* | *MARK1* | - | - | Preserved (11) |
| darkorange | *C6orf118* | *HBG2* | *HBE1* | - | - | New |
| darkred | *CBWD2* | *LRSAM1* | *EVC* | Nicotinate and<br>nicotinamide<br>metabolism | - | Preserved (21) |
| darkturquoise | *PPP1R1C* | *GPR141* | *RASIP1* | - | - | Preserved (8) |
| green | *MAF* | *AGRN* | *LGMN* | - | Dilated<br>cardiomyopathy | Preserved (181) |
| greenyellow | *OPRD1* | *ZBTB2* | *TRIM72* | - | - | Preserved (44) |
| grey60 | *PELI1* | *TEC* | *PDE6A* | Nitrogen metabolism | - | Preserved (20) |
| lightcyan | *LZIC* | *ZER1* | *ZNF670*-<br>*ZNF695* | Valine, leucine and isoleucine degradation | - | Preserved (12) |
| lightgreen | *ARID4A* | *TMEM57* | *DSTYK* | Arginine and proline metabolism | - | New |
| lightyellow | *ANAPC4* | *SCGN* | *KCND3* | - | Various types of N-glycan biosynthesis | New |
| magenta | *AAMDC* | *RYR2* | *GLDN* | - | N-Glycan biosynthesis | Preserved (115) |
| midnightblue | *ULK2* | *OGFOD1* | *SPG11* | - | - | New |
| orange | *CENPI* | *CSMD2* | *TMEM161B* | Pentose phosphate pathway | - | New |
| pink | *DPCD* | *RACK1* | *VWA5B2* | - | Ferroptosis | Preserved (12) |
| purple | *DNMT3A* | *DET1* | *PANK3* | Pantothenate and CoA biosynthesis | - | Preserved (173) |
| red | *JAKMIP1* | *MAP3K12* | *RTCB* | - | - | Preserved (114) |
| royalblue | *RABEPK* | *CCM2* | *GRAMD1C* | - | - | Preserved (9) |
| salmon | *WAS* | *CFHR3* | *SCUBE1* | - | Non-small cell lung cancer | Preserved (17) |
| tan | *DEK* | *AC002310.13* | *AUH* | - | - | Preserved (10) |
| turquoise | *SHISA5* | *RP11*-<br>*164J13.1* | *ARIH2* | Circadian rhythm | Amphetamine addiction | Preserved (151) |
| yellow | *CROT* | *HELB* | *PRELID3A* | Aminoacyl-<br>tRNA<br>biosynthesis | - | Preserved (32) |

**Table 6.** miRNA case network modules, including the top three hub miRNAs in each module as well as enrichment analysis results for top mRNA targets for the top three hub miRNAs in the module. Preservation state shows miRNA cases vs controls preservation analysis results. Modules that were preserved show number of genes preserved between miRNA cases and miRNA controls. Highlighted rows contain miRNAs of interest within the top three eigenmiRs.

| Module | Eigenmirs |  |  | Top KEGG 2021 Pathway<br>for eigenmiR's targeted<br>genes | Preservation<br>State |
| --- | --- | --- | --- | --- | --- |
| blue | hsa-let-7g-5p | hsa-miR-26b-5p | hsa-let-7f-5p | FoxO signaling pathway | Preserved (72) |
| brown | hsa-miR-30e-5p | hsa-miR-191-5p | hsa-miR-199a-3p | Axon Guidance | New |
| green | hsa-miR-378a-3p | hsa-miR-378c | hsa-miR-378d | - | Preserved (29) |
| turquoise | hsa-miR-1268a | hsa-miR-642a-3p | hsa-miR-1268b | - | Preserved (79) |
| yellow | hsa-miR-186-5p | hsa-miR-146a-5p | hsa-miR-221-3p | Notch signalling pathway | Preserved (54) |

Pathway enrichment analysis was then performed using the eigenmiR’s mRNA targets. The following KEGG pathways showed significant enrichment in the brown_(miCases)_ module: long term potentiation, axon guidance, regulation of the actin cytoskeleton, ErbB signalling pathway, T cell receptor signalling pathway, neurotrophin signalling pathway, MAPK signalling pathway, choline metabolism in cancer, FoxO signalling pathway, glioma, cGMP-PKG signalling pathway, renal cell carcinoma, cellular senescence, and B cell receptor signalling pathway. A further enrichment analysis was performed on the top ten eigenmiRs in brown_(miCases)_, within results from the database Jensen COMPARTMENTS, showing significant enrichment of the following entries: extracellular exosome complex, lysosomal multienzyme complex, microvesicle, EmrE multidrug transporter complex, PTEN phosphatase complex, Phosphatidylinositol phosphate phosphatase complex, and apoptotic body.

### Network figures

Modelling of the network with Cytoscape elucidated interesting characteristics within the networks. The mightnightblue(SE) visualised network (Figure 2) contained 12 genes, including the top 10 hub genes from the network, excluding *ELF1*. In addition to the hub genes, the genes *TEX14, ABCB1, and DARS2* appear in the network. An enrichment analysis of these 12 genes showed results for “Carcinoma testes” in the DisGeNET database (adjusted p value < 0.1, OR = 114.02), as well as significant (p value < 0.05) enrichment in several male fertility-related disorders in MGI Mammalian Phenotype Level 4 2021, including male infertility (MP:0001925), decreased testis weight (MP:0004852), and abnormal spermatocyte morphology (MP:0006379).

**Figure 2.**
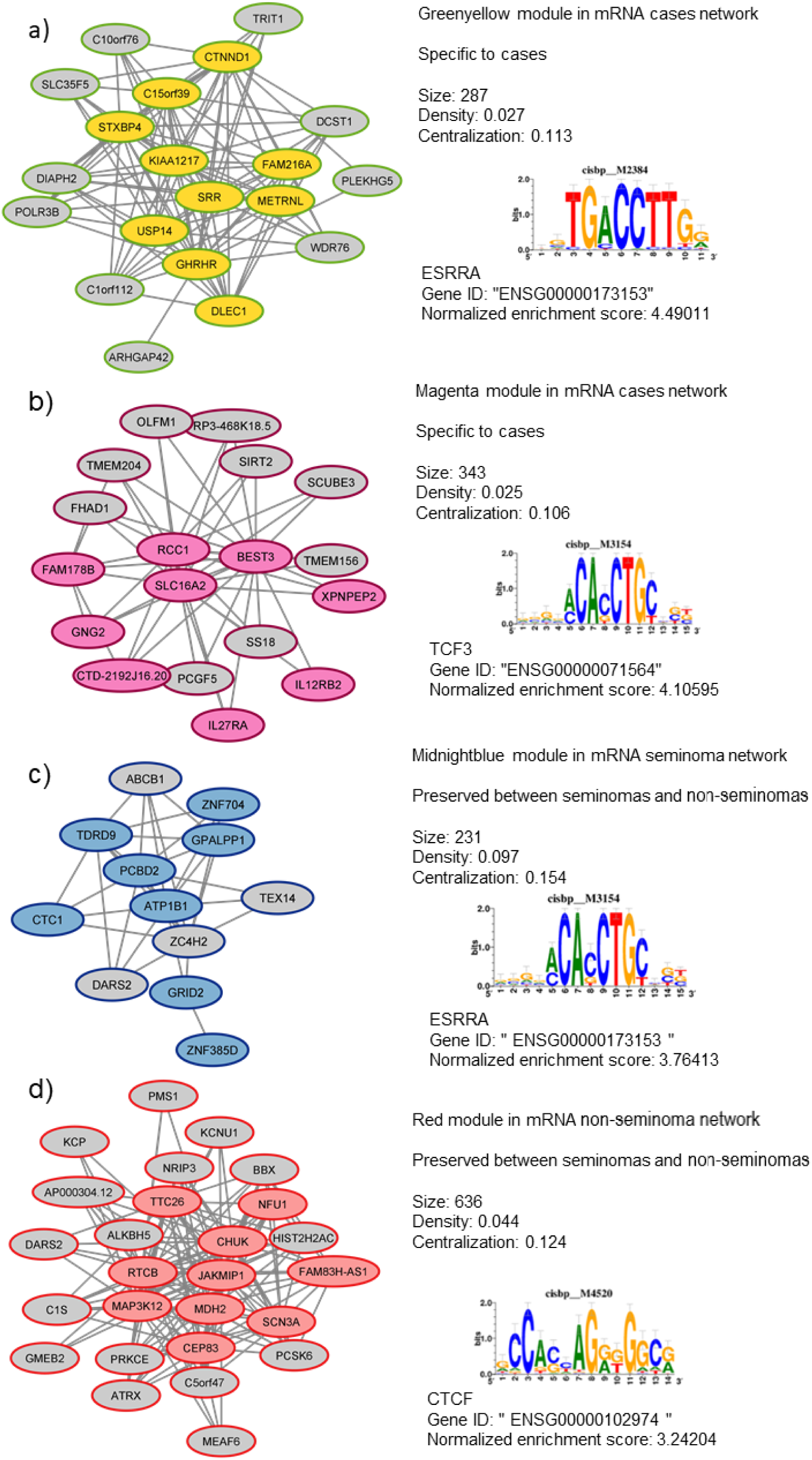
Network model of modules within a,b) mRNA cases, c) seminomas, d) non-seminomas. Connections between genes show a weighted correlation in expression. The distance measurement threshold for this network was set to 0.3. Genes with coloured backgrounds are eigengenes for each module. Sequence motifs show cisbp regulatory element with highest normalised enrichment score for all genes in the module. Module was selected for visualisation due to specificity to TGCT, as well the nodes passing the adjacency threshold set in the analysis.

### Modules with shared eigengenes

Analysis of the eigengenes in modules revealed a pattern emerging in most networks related to TGCT cases. The repeated appearance of the gene MAP3K12 within the eigengenes, and the presence of MAPK signalling pathways within enrichment analysis was further investigated by isolating the modules in which this appeared and determining mRNA commonality between the three. The brown_(miCases)_ module was included through its predicted targets with the same confidence scores used previously. The 21 genes common to all modules were the following: *ATRX, BEND7, DCAF12, FYCO1, GMEB2, GRM3, ICK, IGLON5, KLHL14, MAP3K12, MAPK4, MASTL, NRIP3, PER2, PPP1R37, PRKCE, SCN3A, SLC25A36, STYX, TTLL2, UCHL5* (Figure 3).

**Figure 3.**
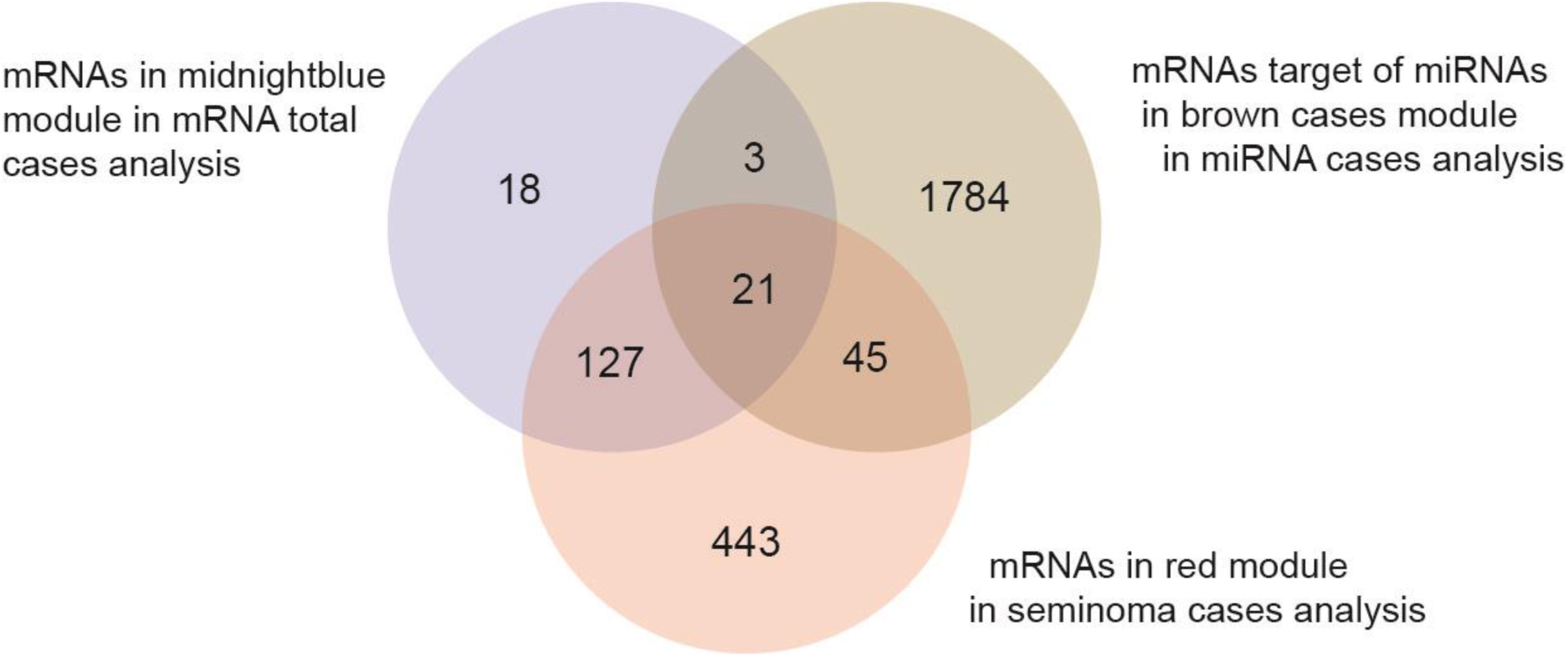
Common genes between each of the three modules containing, either the *MAP3K12* mRNA, or miRNAs that are known to target the *MAP3K12* gene with a confidence score of 70 in miRbase. These modules were midnightblue_(cases)_, brown_(miCases)_ and red_(NS)_. Common genes between these modules were analysed due to the presence of similar eigengenes.

### Regulatory feature prediction

Analysis of the several modules’ genes using the prediction tool i-cis Target showed regulatory elements with possible links to TGCT development. For both the modules greenyellow_(cases)_ and midnightblue_(SE)_, the highest scoring regulatory elements from the cisbp database was oestrogen-related receptor alpha gene transcription factor (*ESRRA*), with *ESRRG* also amongst the highest scoring for midnightblue_(SE)_ (Figure 3). The red_(NS)_ module’s highest scoring regulatory element was associated with *CTCF*, and for the magenta(_cases_) module, the highest score regulatory feature was associated with *TCF3*.

## Discussion

In our previous analysis of RNA profiles from the Janus Serum Bank samples using linear models we identified RNAs which were differentially expressed in patients prior to diagnosis of TGCT. These included genes such as *TEX101,* a male fertility-related gene, and *TSIX,* X-chromosome inactivation genes (18, 41). As gene products and regulatory elements often operate in close relationships, we used network analysis framework and the tool WGCNA to identify interplay between genes and ncRNAs that could be involved in TGCT development in the years leading up to diagnosis. We combined miRNA and mRNA analyses by using miRNA targets, to further validate both types of RNA. When comparing the networks constructed from mRNA case and mRNA control data, we were able to identify several modules specific to cases that contained genes that were previously known to be associated with TGCT.

### mRNA Cases Network

Within the mRNA cases network there were several modules with eigengenes associated with both TGCT and male subfertility. The new black_(cases)_ module’s top eigengene, *UBCA1*, has been previously seen to be downregulated in TGCT tumour samples compared to controls (42). This suggest that *UBCA1* is involved in early TGCT development and could be used as a potential biomarker. In addition, links to early defects in the male reproductive system of mice have been described with *UBE2W*, the second hub gene in this module (43). In the study, knock-out mice were used to investigate the role UBE2W plays in testis development. It was observed that the testis is vulnerable to the loss of UBE2W as seen through the high rate of male infertility after loss. Alongside this, UBE2W knock-out causes an increase in testicular vacuolation defects, pointing to its role in the maintenance of the testis structure. Defects in the male reproductive system of humans were also linked to the third hub gene, *OSA3*. Missense variants of the *OSA3* gene were found in members of a 22-family group with recurrent disorders of sexual development and hypospadias, a component of the testicular dysgenesis syndrome, often associated with TGCT (44). *UBE2W* and *OSA3* were also present in the blue_(SE)_ module’s eigengenes, and *UBCA1*, *UBE2W,* and *OSA3* appeared in the red_(NS)_ module’s eigengenes. This indicates that these genes may play an important role in the formation of both seminomas and non-seminomas due to their repeated appearance as eigengenes in both the overall cases, and in both histological types.

Within the new magenta_(cases)_ module, the eigengene *RCC1* has known associations with TGCT. *RCC1* can form a fusion gene with *ABHD12B,* and this fusion, *RCC1-ABHD12B,* has been shown to be present in 9% of TGCT tissues but was not present in any control testis tissues (45). However, another study showed expression of *RCC1-ABHD12B* in 60% of GCNIS samples, in 80% of seminoma samples, and in all embryonal carcinoma cell lines (46). In this study, another fusion, *RCC1-HENMT1,* was also observed in a significant number of TGCT tissues of undifferentiated histological subtypes of TGCT, including all GCNIS samples, and all other TGCT samples, excluding on yolk sac tumour.

The MAPK signalling pathway-related gene *MAP3K12*, was observed as an eigengene for the new midnightblue_(cases)_ module. Previously *MAP3K12* has been associated with prostate cancer, and regulation of *MAP3K12* by miRNAs could suppress prostate cancer progression (47). *MAP3K12* was also observed as one of the genes associated with activation of oncogene-induced angiogenesis in hepatocellular carcinomas (48). MAPK signalling pathways have also been labelled as one of the dominant functional pathways in spermatogenesis, alongside the AMPK pathways (49). We observed both MAPK-related genes and significant association with AMPK-related pathways in the new modules of the mRNA cases network. The involvement of these pathways in TGCT development should be investigated further as they may play an important role.

In addition, the new purple_(cases)_ module eigengene *FMR1* has been investigated in TGCT patients and could be used to determine overall prognosis (50). It was observed that expression levels of *FMR1* were positively correlated with the clinical outcome of TGCT and therefore could act as tumour suppressors alongside *AR* and *GPC3* genes (50). The eigengene, *RBFOX2*, has been previously studied in cryptorchidism susceptibility. Paralogs of this gene are expressed in the gubernaculum; failure of this fibrous cord to elongate during development causes the birth defect commonly associated with TGCT (51, 52).

The new Salmon_(cases)_ module contained the eigengene *IGF2BP3*, also known as *IMP3*. The IMP3 protein is an oncofoetal protein which has known roles in both embryogenesis and carcinogenesis (53). Within teratomas, IMP3 expression has been seen in 100% of mature teratoma components and in 96% of all metastatic testicular teratomas. In this study, it was also observed to be expressed in 99% of all other TGCT components (53). Therefore, it should also be investigated further for possible role in development of GCNIS and its progression to TGCT, due to its shared roles in embryogenesis and carcinogenesis.

### Histology-Related Networks

Within the seminoma network, one of the modules, the new lightyellow_(SE)_ module, contained the eigengene *G3BP2*. Studies into TGCT have identified the cytogenetic band 4q21.1 as being a possible location for a susceptibility locus, the genes positioned within this band include *G3BP2,* whilst not fully labelled as a TGCT susceptibility gene, its location within this band should be noted (8). Our observation of this gene within seminomas could explain its identification within genome-wide associations studies (GWAS), and its role within seminoma histology should be investigated further.

The preserved midnightblue module_(SE)_ also contained an eigengene that has previously been identified in TGCT studies (Figure 2). The *GRID2* gene is an extremely large gene (1.47Mbs within 4q22.3) that could be involved in hepatocellular carcinomas (54). Focal deletions in *GRID2* have been seen in TGCT, though it has primarily been seen within non-seminomas (55). Finally, within the non-seminoma networks, the module yellow_(NS)_ contained the eigengene *NARS2,* which has previously been identified as a TGCT susceptibility gene through GWAS within the cytogenetic band 11q14.1 (8), and is also involved in ovarian serous cystadenocarcinomas as a possible progression associated gene (56).

Eigengenes related to TGCT, subfertility, and disorders of sexual development in the case networks are potential liquid biopsy biomarkers. With samples being taken up to 10 years before diagnosis and still exhibiting male reproductive health-related genes, it could be beneficial to look more closely into the pathways associated with these RNAs and the reason for their presence in serum. Although there is not one common pathway or mechanism between these eigengenes, we know that TGCT and cancer in general are multifaceted and complex. Even within histological subtypes of TGCT, tumour growth stems from multiple cell types with multiple mechanisms, and therefore we should expect many shared pathways for all the eigengenes related to TGCT.

TGCT analysis using network methods has been performed previously on TGCT tissue, which has allowed the identification of genes that are differentially expressed between histological subtypes (31). In particular, the transcription factor-target gene pairs, *MYC* and *SP1,* were found to be common eigengenes in both seminomas and non-seminomas, as well as the miRNA, miR-182-5p (31). *MYC* have an oncogenic association with TGCT (57), *SP1* is thought to be involved in the process of aerobic glycolysis in TGCT cells (58), and miR-182-5p and its targets are associated with cancer cell proliferation and migration (59, 60). However, within our pre-diagnostic data, we did not see *MYC* or *SP1* as eigengenes in any of the seminoma or non-seminoma modules. This could mean that these genes are not associated with progression of GCNIS to TGCT but could be involved in the maintenance of the malignancies via the previously described processes.

### Inter-network features

The mRNAs, *RTCB*, *JAKMIP1,* and *MAP3K12*, as well as the miRNA hsa-miR-30e-5p were present as eigengenes in nearly all case related modules, and hsa-miR-30e-5p is thought to potentially target both *MAP3K12* and *RTCB*. When investigating the common mRNAs shared between all three, the presence of fertility- and cancer-related genes was observed. Early spermatid production relies on ion channels for both overall fertility and normal sperm physiology, and the gene *SCNA3* is involved in this process and defects within these channels are associated with male infertility (61). The precise role of *ATRX* in human gonadal development is still not entirely clear, however, links between the gene *ATRX* and *DMRT1* have been previously noted. It is also thought that ATRX acts downstream of the genes *SPY* and *SOX9*, which are crucial for Sertoli cell development and upregulation of anti-Müllerian hormone during testicular development *in utero* (62, 63). Within mice models, *Styx* has been seen to be strongly involved in spermatid development, with ablation of *Styx* in mice causing a 1,000-fold reduction in spermatozoa production (64). These shared genes between three case-specific modules show common male fertility-related associations, indicating again the presence of possible markers for TGCT progression.

Another shared component between networks was the midnightblue_(SE)_ module. When comparing the seminoma and non-seminoma networks together, it was noted that this midnightblue module was significantly preserved between the two histological states. Modules that were shared between seminomas and non-seminomas could offer insights into common pathways and genes of interest common to most types of TGCT. Initially the gene GRID2 had been identified as a gene of interest due to its associations with non-seminomas. However, when visualising the preserved midnightblue_(SE)_ module in Cytoscape (Figure 2), further genes of interest were noted, such as the gene *TEX14,* which was associated with the hub genes for the module, though not an eigengene itself. *TEX14* has been identified as a susceptibility locus for TGCT (8). As shown previously, other TGCT susceptibility genes also appeared amongst the hub genes of modules in both the seminoma and non-seminoma modules, including *NARS2* in the yellow_(NS)_ module and *G3BP2* in the lightyellow_(SE)_ module. Both these modules are significantly preserved between both histological states. Our findings are in line with the GWAS identifying these loci regardless of histology, however, our findings propose that changes at these loci may be observable earlier than previously thought.

In the midnightblue_(SE)_ and the greenyellow_(cases)_ modules, we also saw the enrichment of EGRR genes during the regulatory elements analysis. *ERR* are a group of genes that all code for oestrogen-related receptor proteins, and previously they have been found to be crucial in controlling energy metabolism for both normal and for cancerous cells, particularly ERRA and ERRG (65). In some cancers, such as endometrial, colorectal, and prostate cancer, ERR expression is low, which suggests that ERRs have an overall negative effect on tumour progression (66, 67). In the formation of Leydig cells, ERRs are also crucial, as lack of ERRs has been shown to affect Leydig cell morphology and physiology in cultures (68). ERRA has also shown to mediate the adaption of metabolic systems to the tumour microenvironment and could be involved in the direct activation of Wnt11/β-catenin pathway, and this in turn leads to an increase in the capacity of cells to migrate (68). Overall, these genes are strongly involved in the molecular characteristics of Leydig cell tumours. Although this is a less common form of TGCT, the presence of ERR transcription factor enrichment in our seminoma samples could indicate a joint aetiology between the two tumour types.

Further analysis of regulatory elements identified transcription factors for the gene *CTCF* as being associated with the genes within the red_(NS)_ module. CTCF and its paralogues have been previously identified in testicular tumour cells. It has been suggested that the co-expression of CTCF and BORIS could be responsible for epigenetic deregulation leading to cancer. Furthermore, the mechanisms surrounding CTCF/BORIS could be an essential factor leading to the immortalisation of testicular cancer cells (69).

### Strengths and Limitations

The strength of WGCNA is the reduction in the complexity of the data, allowing for a more accurate interpretation. With this approach we have detected TGCT-related genes in addition to those previously identified with linear models. The size and scope of the dataset is also advantageous and allows us to investigate pre-diagnostic TGCT patients, whilst also considering confounding factors from health surveys.

However, sample size is still a limitation with this study. Within this dataset, there is a distribution amongst different RNA types, whilst we were able to perform a network analysis for different histological types on the mRNA from our data, the scarcity of other RNA types limited sub-analysis on seminoma and non-seminoma for other types, including miRNA. The mRNA levels were derived from small RNA sequencing of long-term stored serum samples. We assume the short mRNA fragments, up to 47 nucleotides in length, represent the full-length mRNA profiles. An increase in statistical power would benefit the WGCNA analyses across histological types as well as alternate RNA types.

### Conclusion

The primary insights gained from this work include multiple pathways and genes with possible associations to TGCT initiation or growth, providing better understanding of the development of this disease. We see TGCT-related genes with high degrees of connectivity in TGCT exclusive networks, such as *TEX14*, *NARS2*, and *G3BP2*. In addition, we observe the enrichment of cancer-related mismatch repair KEGG pathway and have identified predicted transcriptional factors such as oestrogen-related receptors (ERR) in multiple TGCT modules. Enrichment analysis also showed “Carcinoma testes” and several male fertility-related disorders as significantly enriched within a seminoma module.

Furthermore, there is the potential for RNAs discussed here to be utilised as biomarkers for earlier detection. Possible future studies could involve the use of knock-out cell lines, including 3D models (70), for eigengenes discovered in the modules of our networks. Systematic investigation of these genes and their roles in cell lines would allow for validation of the altered phenotypes for each of these genes and propose a possible order of molecular events in tumorigenesis.

## Data Availability

The datasets generated for this article are not readily available because of the principles and conditions set out in articles 6 (1) (e) and 9 (2) (j) of the General Data Protection Regulation (GDPR). National legal basis as per the Regulations on population-based health surveys and ethical approval from the Norwegian Regional Committee for Medical and Health Research Ethics (REC) are also required. Requests to access the datasets should be directed to the corresponding authors.

## Author Contributions

MW conceptualized the network studies. All authors designed the study. JB and MW performed the analyses of the data. All authors drafted the manuscript. All authors interpreted and discussed the results, contributed to the writing, and approved the final manuscript.

## Acknowledgements

We would like to acknowledge Sinan U. Umu for his contribution with the pre-processed analysis and management of the Janus Serum Bank sncRNA data.

## Ethics Statement

The studies involving human participants were reviewed and approved by the Norwegian Regional Committee for Medical and Health Research Ethics (REC no: 2016/1290). The patients/participants provide their written informed consent to participate in this study.

## Online Appendix

### Appendix 1

#### Sensitivity Analysis

Sensitivity analysis of the mRNA networks was performed through the exclusion of samples diagnosed from 8-10 years after sampling. Weighted correlation gene networks were constructed using the remaining samples diagnosed from 0-7 years after sampling with matched controls. For the sensitivity analysis of 0-7 years, the cases network consisted of 26 modules with an average module size of 412. The largest module, turquoise_(cases 0-7)_ consisted of 1948 mRNAs and had a density of 0.012 and a heterogeneity of 0.529. Control network with 8–10-year samples excluded consisted of a total of 14 modules with an average module size of 765. The largest module here was turquoise_(controls 0-7)_ which consisted of 2200 mRNAs with a density of 0.033 and a heterogeneity of 1.324.

Enrichment analysis of 0-7 years showed several new modules in both cases and control networks. The turquoise_(cases 0-7)_ module finds both cancer-related and germ cell development-related pathways during enrichment analysis.

Further sensitivity analysis was performed through the exclusion of all samples diagnosed 6-10 years after sampling. Weighted gene correlation networks showed that using 0–5-year samples would allow for the construction of a network with 28 modules in total. The average module size was 382.

**Supplementary Figure 1.**
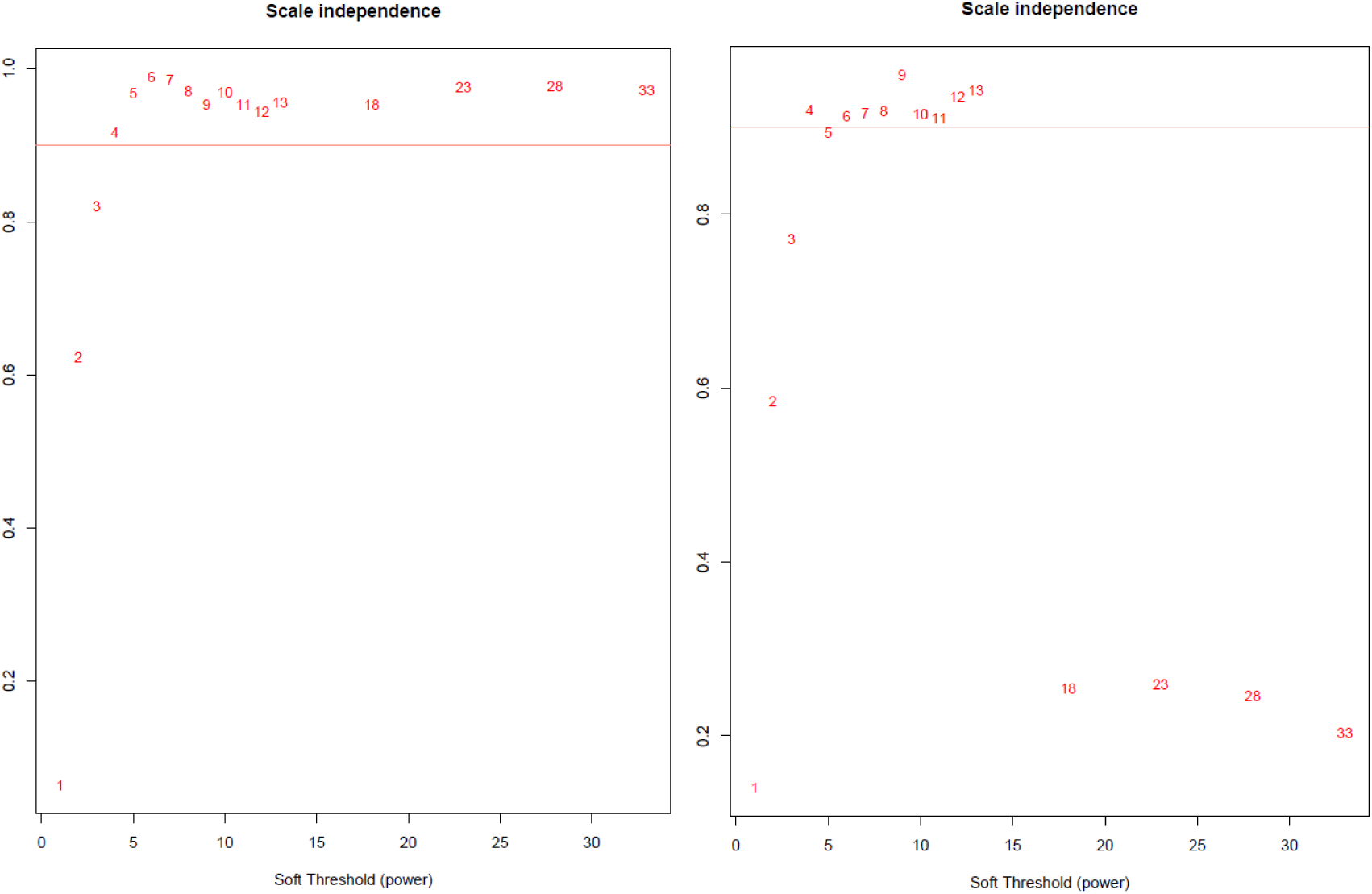
Power analysis for mRNA (A) and miRNA (B), showing 10 as the appropriate soft threshold for weighted gene correlation network analysis.

